# The Impact of Scaling up Dolutegravir on Antiretroviral Resistance in South Africa

**DOI:** 10.1101/19010132

**Authors:** Anthony Hauser, Katharina Kusejko, Leigh F. Johnson, Huldrych F. Günthard, Julien Riou, Gilles Wandeler, Matthias Egger, Roger D. Kouyos

## Abstract

**Background:** Rising resistance of HIV-1 to non-nucleoside reverse transcriptase inhibitors (NNRTIs) threatens the success of the global scale-up of antiretroviral therapy (ART). The switch to WHO-recommended dolutegravir (DTG)-based regimens could reduce this threat due to DTG’s high genetic barrier to resistance. We used mathematical modelling to examine the impact of the scale-up of DTG-based ART on NNRTI pre-treatment drug resistance (PDR) in South Africa, 2019-2040.

**Methods and results:** We adapted the MARISA (Modelling Antiretroviral drug Resistance In South Africa) model, an epidemiological model of the transmission of NNRTI resistance in South Africa. We modelled the introduction of DTG in 2019 under two scenarios: DTG as first-line regimen for ART-initiators, or DTG for all patients, including patients on suppressive NNRTI-based ART. Due to safety concerns related to DTG during pregnancy, we assessed the impact of prescribing DTG to all men and in addition to i) women beyond reproductive age, ii) women beyond reproductive age or using contraception, and iii) all women. The model projections show that, compared to the continuation of NNRTI-based ART, introducing DTG would lead to a reduction in NNRTI resistance in all scenarios if both ART initiators are started on a DTG-based regimens and those on NNRTI-based regimens are rapidly switched to DTG. NNRTI resistance would continue to increase if DTG-based ART was restricted to men. When given to all men and women, DTG-based ART could reduce the level of NNRTI resistance from 58.5% (without DTG) to 14.8% (with universal DTG) in 2040. If all men and women beyond reproductive age or on contraception are started on or switched to DTG-based ART, NNRTI resistance would reach 35.1% in 2040.

**Conclusions:** Our model shows the potential benefit of scaling up DTG-based regimens for halting the rise of NNRTI resistance. Starting or switching all men and women to DTG would lead to a sustained decline in resistance levels whereas using DTG-based ART in all men, or in men and women beyond childbearing age, would slow down the increase in levels of NNRTI resistance.

## Introduction

The rollout of antiretroviral therapy in South Africa is estimated to have prevented 0.73 million HIV infections between 2004 and 2013 as well as 1.72 million deaths between 2000 and 2014 [1, 2]. However, the spread of non-nucleoside reverse transcriptase inhibitor (NNRTI) resistant viruses is threatening this success [3]. An estimated 16% of AIDS-related deaths and 8% of ART costs will be attributable to HIV drug resistance up to 2030 in the sub-Saharan African countries that reached HIV pre-treatment drug resistance levels above 10% in 2016 [4].

In Southern Africa, dolutegravir (DTG), an integrase inhibitor drug, will be introduced on a large scale as part of fixed-dose combinations of Tenofovir, Lamivudine, and Dolutegravir (TLD) [5]. With a high genetic barrier to resistance, DTG has the potential to curb the spread of antiretroviral resistance, as it is highly effective, well tolerated and affordable in resource-limited settings [6, 7, 8, 9]. Mathematical models explored the effectiveness and cost-effectiveness of prescribing DTG to all ART-initiators [10]. These models found that the introduction of DTG was cost-saving and reduced HIV mortality in people living with HIV who initiate ART [10]. The introduction of DTG has been complicated by the increased risk of neural tube defects (NTD) in women living with HIV using DTG at the time of conception [11] and other potential side effects such as weight gain [12, 9J]. Concerns surrounding NTD risk have delayed the rollout of DTG and, in some settings, led to recommending DTG-based regimens only for men and women who are not at risk of pregnancy [13, 14]. For South Africa, a mathematical modelling study showed that DTG-based first-line ART for all women of child-bearing potential would prevent more deaths among women and more sexual HIV transmissions than either NNRTI-based ART for women of child-bearing potential or women without contraception, but increase pediatric deaths [15]. In its 2019 guidelines, the WHO recommends DTG in combination with nucleoside reverse-transcriptase inhibitors (NRTI) for first-line ART, with the proviso that “women should be provided with information about benefits and risks to make an informed choice regarding the use of DTG” [16].

It is likely that in many settings people living with HIV on NNRTI-based first-line ART will be switched to DTG-based ART, however, the rate of the transition will vary between countries and settings. For second-line ART, WHO recommends DTG-based ART in people living with HIV for whom a NNRTI-based first-line regimen has failed [16]. Again, the rate of switching to DTG-based second-line ART will vary, influenced by concerns about the development of dolutegravir resistance in patients who switch with pre-existing resistance to NRTIs [17]. Taken together, it is likely that for the foreseeable future a considerable fraction of people living with HIV, and particularly women, may continue to rely on NNRTI-based ART regimens, even in the case when guidelines recommend DTG. In this context, NNRTI resistance will likely remain an important issue during and after the rollout of DTG.

We adapted the MARISA model (Modelling Antiretroviral drug Resistance In South Africa) [18] to examine the impact of different scenarios regarding the scale-up of DTG-based ART on NNRTI pre-treatment drug resistance (“NNRTI resistance” in the remaining of this article) in South Africa for 2019-2040.

## Materials and methods

### Extended MARISA model

Described in detail elsewhere [18], MARISA is a deterministic compartmental model of both the general HIV epidemic and the NNRTI resistance epidemic in South Africa. It consists of four dimensions representing i) care stages (see Fig 1), ii) disease progression according to the CD4 cell counts, iii) NNRTI resistance, and iv) gender. Care stages distinguish between infected, diagnosed and treated individuals (either with NNRTI- or protease inhibitor (I)-based regimen), with subsequent treatment-specific suppression (*Supp* compartment in Fig 1) or failure (*Fail* compartment). *Treat init*. compartments represent individuals treated for less than 3 months.

**Figure 1.**
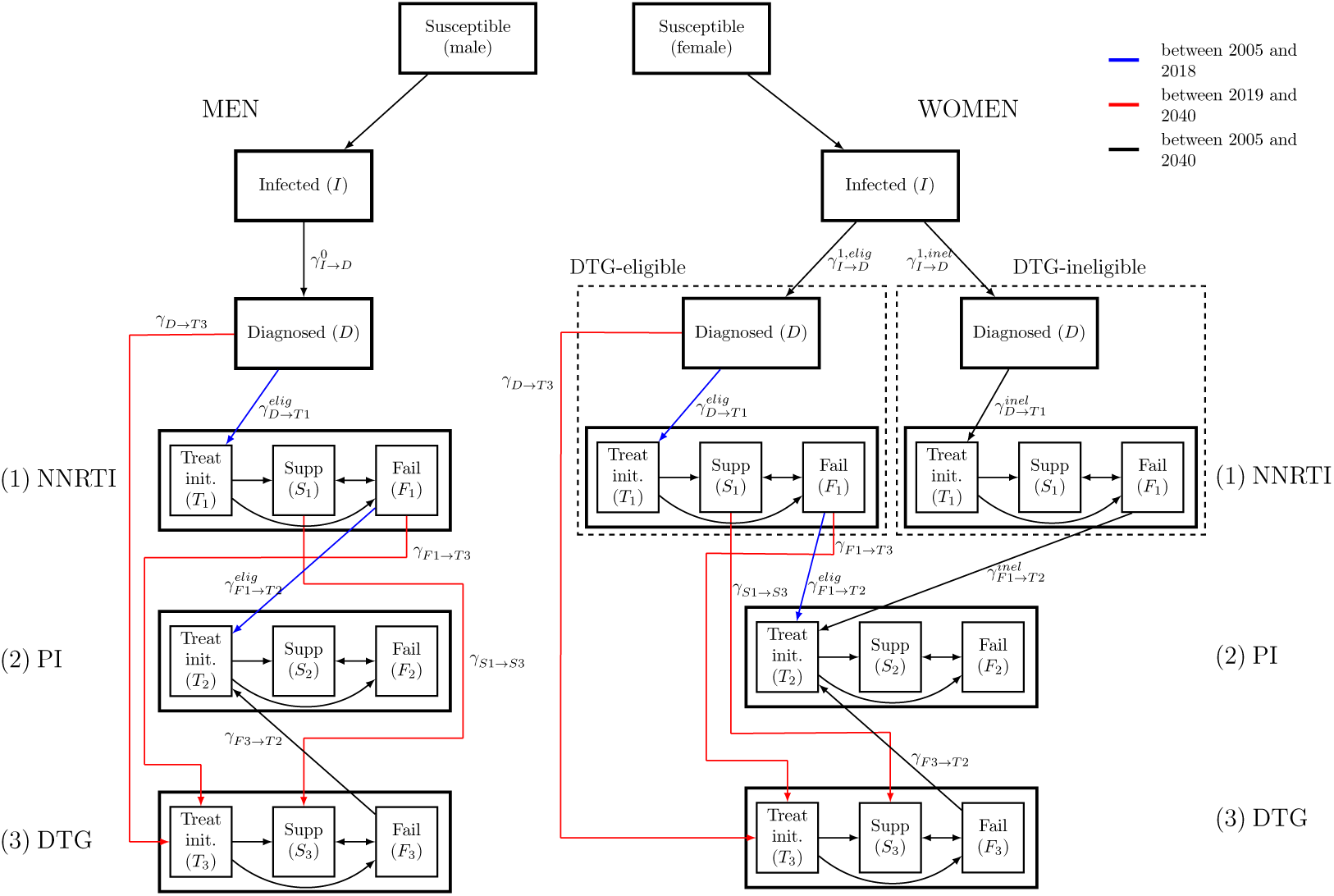
The adapted MARISA model. The model differentiates DTG-eligible from DTG-ineligible women. The model structure related to the cascade of care is shown.

We simulated the adapted model from 2005 to 2040 assuming the rollout of DTG-based ART started in 2019 under different scenarios (see below). We further assumed that all men and a proportion *p*1 of women are eligible for DTG, this proportion varied between scenarios. Before 2019, an NNRTI is used in first-line ART and PIs in second-line regimens. As first line regimen, DTG is prescribed from 2019 on either to ART initiators (i.e. eligible, ART-naive people living with HIV) or for switching to DTG-based first-line ART (i.e. people on NNRTI and eligible for DTG). We assumed that patients failing DTG are switched to a PI-based regimen. For DTG-ineligible women, the cascade of care remains unchanged after 2019 (Fig 1).

### Calibration and extension of the MARISA model

We previously calibrated the MARISA model by combining different sources of data. Rates either related to treatment response (NNRTI- or PI-based regimen) or disease progression (characteried by CD4 counts) were estimated using clinical data from data from five cohorts in South Africa (Aurum Institute, Hlabisa, Khayelitsha, Kheth Impilo and Tygerberg) that participate in the IeDEA collaboration [19]. These cohorts provided longitudinal information for 54,016 HIV-infected adults. Other parameters were either estimated from the literature (e.g. resistance-related parameters) or fitted to estimates from the Thembisa model (e.g. diagnosis rates and treatment initiation rates). Thembisa is a demographic projection model on which the official UNAIDS estimates for South Africa are based [20]. More details about the calibration procedure can be found in [18] and in the supplementary materials (S1 Text, Section 1.2).

We added and modified parameters in order to model the introduction of DTG. We assumed that the DTG initiation rate *γ*_*D→T* 3_(*t*) is the same as the NNRTI initiation rate *γ*_*D→T* 1_(*t*) from 2019. Both NNRTI and DTG initiation rates increase until 2022, as a consequence of the Treat-All policy that was implemented in 2017. From 2022 onwards, they are assumed to remain constant (S1 Text, Section 2.1). Finally, we fixed switching rates from NNRTI-to DTG-based regimens for both eligible suppressed (see “Scenarios”) and all failing individuals (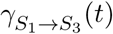 and 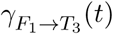 respectively) to 1 year^*−*1^. Finally, we assumed that suppressed individuals would stay suppressed when switching, while failing individuals would start DTG in the Treat init. compartment (see Fig 1). The main parameters are summarized in Table 1.

**Table 1.**
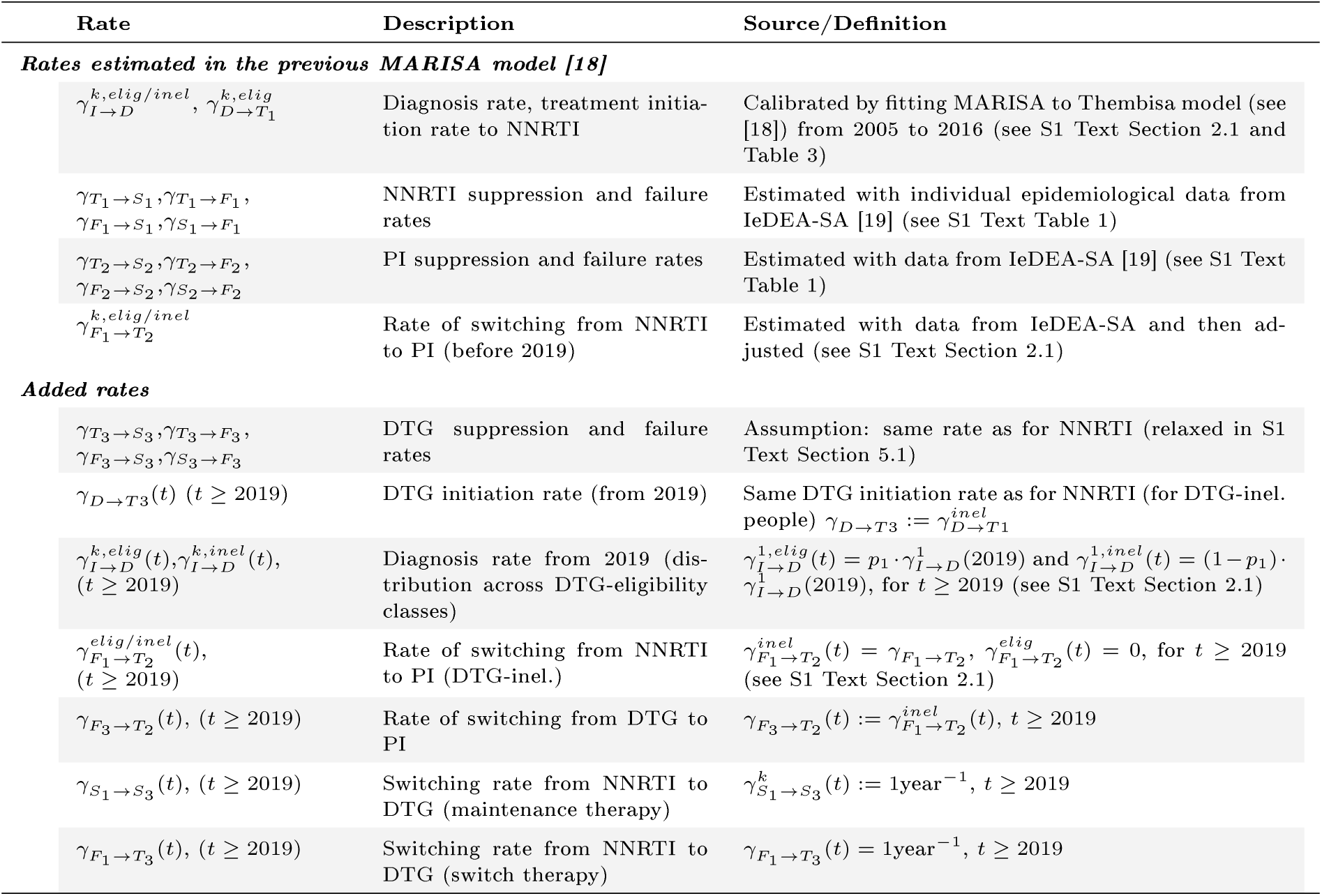
Main parameters used in the extended MARISA model.

### Scenarios

The model investigated the impact of the introduction of DTG-based regimens on the level of NNRTI resistance in diagnosed individuals under two main scenarios, with four variations for each of the two scenarios. We also examined the scenario where DTG-based ART is not introduced. There were thus nine scenarios in total. The two main scenarios were:

1. DTG only used in first-line regimen of ART-initiators and, as second-line, in patients failing NNRTI-based ART,
2. DTG used as initial first-line regimen (for ART-initiators), with all patients on NNRTI-based regimens being switched to a DTG-based regimen.

Each scenario also investigated four different DTG eligibility levels *p*1 for women. The population eligible for DTG in each scenario was:

a. only men (100% men, 0% women)
b. men and women beyond reproductive age (100% of men, 17.5% of women)
c. men and women beyond reproductive age or using contraception (100% of men, 63% of women)
d. all men and women (100% of men, 100% of women)

The percentages of women eligible for DTG in b) and c) were determined by analyzing cohort data from IeDEA, which show that 17.5% adult women on ART are 50 or older [19], and estimates on the use of contraception from the World Bank [21] (see S1 Text, Section 3.1). Of note, to model scenario 1., we set 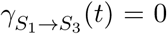 and 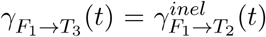 (as opposed to scenario 2., where 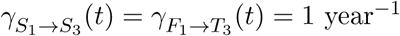). We thus assumed that in all scenarios DTG will be used in second-line regimens for people failing NNRTI-based first-line ART.

### Additional analyses

We predicted the impact of different levels of DTG introduction on the level of NNRTI failure. We considered the scenario where DTG was prescribed to ART initiators and those on NNRTI-based first-line ART were switched to a DTG-based regimen, with the four different levels of women accessing DTG-based ART (see “Scenarios”). However, we assumed that 99% of women were eligible for DTG in scenario d) (instead of 100%), in order to estimate NNRTI failure when only a very small fraction of women rely on it. For each of these scenarios, we predicted the percentage of individuals failing an NNRTI-based regimen in different calendar years (2025 and 2035) and after different durations on ART (1 or 2 years). For each of the scenarios, we ran the model from 2005 up to 2035 and retained the numbers of people starting NNRTI-based first-line ART (by CD4 groups, NNRTI-resistance and gender) in 2025 and 2035. We then ran the model for the compartments related to NNRTI-treatment, using the previously saved starting values. This way, we could predict the levels of NNRTI-failure in patients starting NNRTI in either 2025 or 2035 after 1 or 2 years of ART.

We assessed the impact of different switching rates from NNRTI-to DTG-based regimens, fixed to 1 year^*−*1^ for both suppressed and failing individuals. We varied both rates 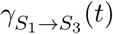 and 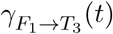 within a range corresponding to a time to switch of between 0.5 and 10 years after start of ART. For each analysis, the percentage of women who are DTG-eligible varied from 0% to 100%.

### Sensitivity analyses

The values of eight parameters were varied in the sensitivity analysis: three transmission-related parameters (percentage of men who have sex with men (MSM), probability of male-to-male infection per sexual contact, and HIV prevalence ratio between MSM and heterosexuals), four resistance-related parameters (resistance rates, reversion to wild-type rate and the effect of NNRTI-resistance on NNRTI efficacy) and one parameter related to treatment (efficacy of DTG-based treatment). Multivariate uncertainty within specified ranges was assessed using Latin hypercube sampling [22]. Each model estimate is reported with a 95% sensitivity range. Further details are available in S1 Text, Section 3.2. In addition, we also investigated the impact of lower treatment-initiation rates than suggested by the Treat-All policy (as suggested by [23], treatment interruption and higher efficacy of DTG on NNRTI PDR (S1 Text, Section 5).

## Results

### Use of NNRTIs and levels of resistance

The percentages of patients treated with NNRTI and PI for each of the nine scenarios are shown in Fig 2. The predicted evolution of levels of NNRTI resistance up to 2040 across the nine scenarios is shown in Fig 3. The model predicts that while NNRTI resistance would increase substantially under continued NNRTI-based ART, the introduction of DTG-based ART can halt this increase, if in addition to starting new patients on a DTG-based regimen the patients on NNRTI-based regimens are switched to DTG-based first-line ART. Specifically, under the scenario of continued NNRTI-based ART as standard first-line therapy, NNRTI resistance would increase to 46.8% (95% sensitivity range: 19.7%-54.5%) by 2030 and 58.5% (32.5%-68.9%) by 2040 (Fig 3A). At the other end of the spectrum, initiating all new ART patients on DTG-based ART and rapidly switching all patients currently on NNRTI-based ART to DTG-based regimens, independently of their gender, would stabilize NNRTI resistance at a low level, with a prevalence of 14.3% (3.5%-17.5%) by 2030 and 14.8% (6.6%-19.5%) by 2040 (Fig 3B). Using DTG only in first-line regimens of patients initiating ART is not sufficient to curb the increase of NNRTI resistance, even when given to all men and women (Fig 3A).

**Figure 2.**
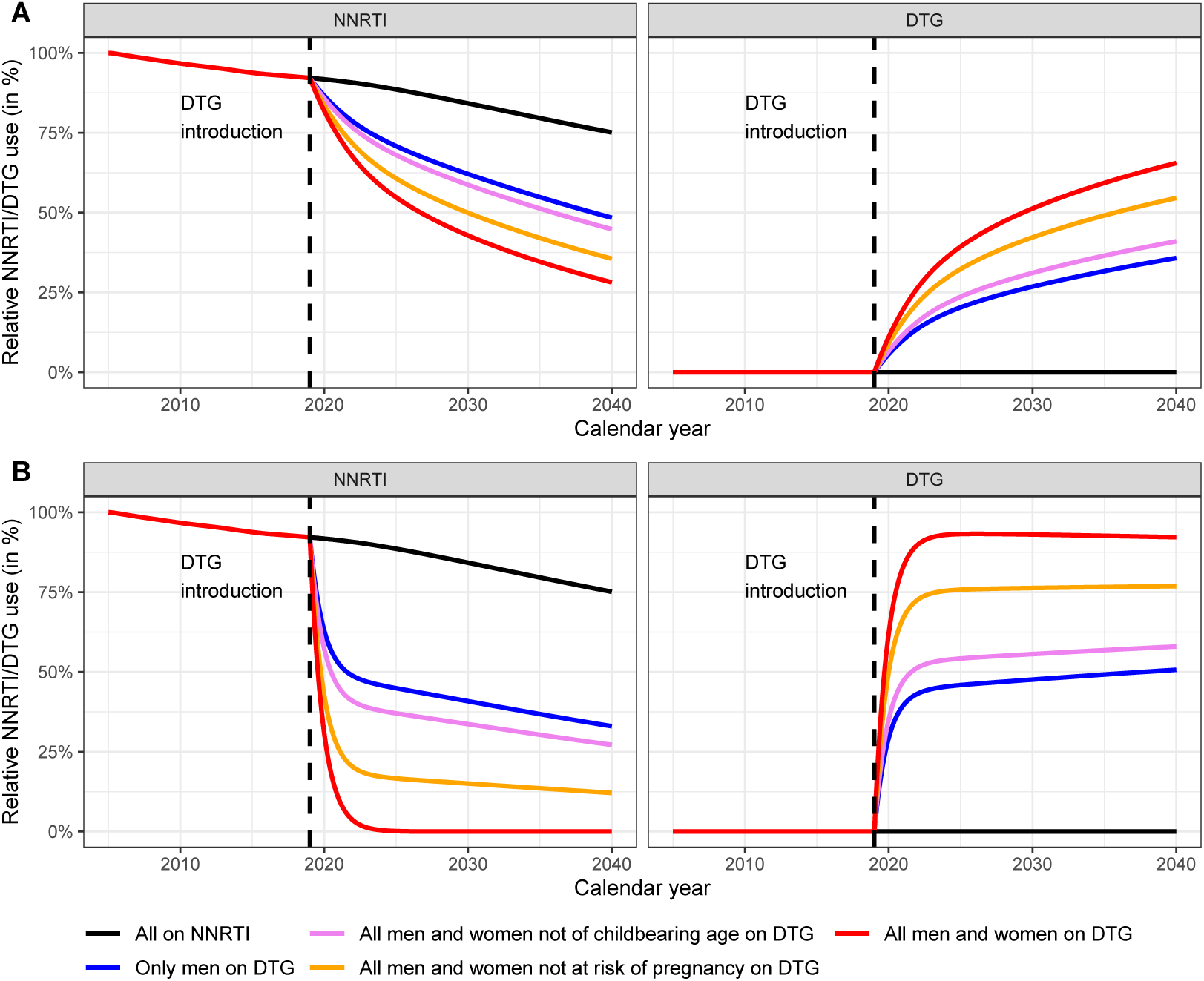
Predicted use of NNRTI- and DTG-based regimens. Percentages of patients treated with NNRTI- and DTG-based regimens (left and right panels, respectively) are shown. anels A represent the scenarios where DTG is used in patients initiating ART, while in panels B patients are also switched to DTG-based first line ART.

**Figure 3.**
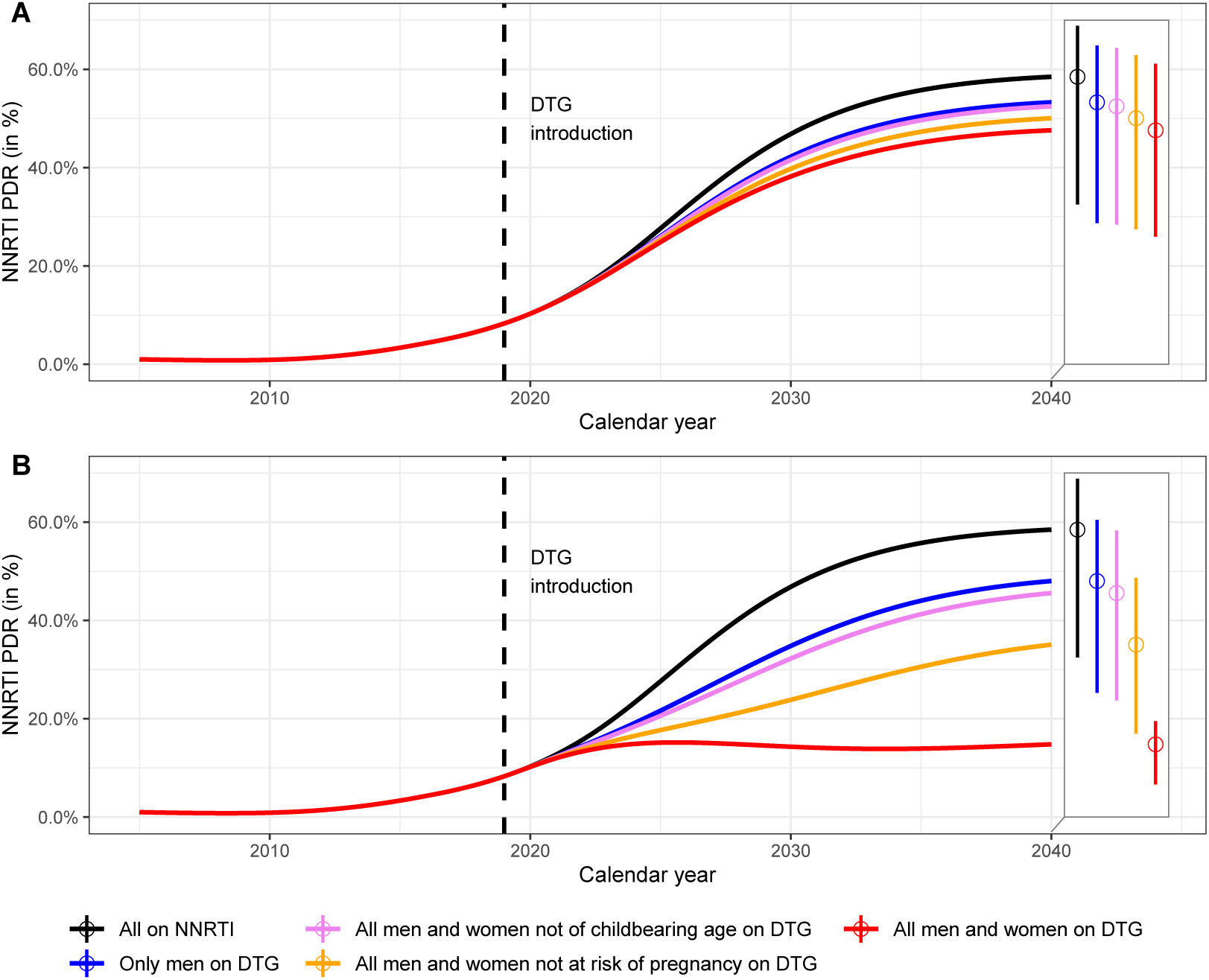
Predicted levels of NNRTI pre-treatment drug resistance in South Africa (PDR) 2005-2040. Dolutegravir is introduced in 2019 under two scenarios: DTG as first-line regimen for ART-initiators (panel A) or DTG for all patients (panel B), and with different eligibility criteria for women. The baseline model shows the situation without the introduction of dolutegravir (black line). The two boxes on the right of each panel represent the levels of NNRTI resistance in 2040 and their 95% sensitivity ranges.

Restricting DTG-based ART to men to avoid the risk of DTG-associated neural tube defects in newborns will not curb the increase in NNRTI resistance: the prevalence of resistance is predicted to increase over the entire study period, reaching values of close to 50% by 2040 (Fig 3B). The situation is similar under the scenario of initiating or switching men and women beyond childbearing age (17.5% of women in the IeDEA cohorts). However, the model estimates that the increase in the prevalence of NNRTI resistance is substantially slowed down if women beyond the age of reproduction or on contraception (63% of women) also initiate a DTG-based regimen or switch to DTG. Under this scenario the prevalence of NNRTI resistance is predicted to reach 35.1% (17.0%-48.7%) in 2040 (and 23.9% (7.7%-29.3%) by 2030) if DTG is given to both ART-initiators and individuals already on NNRTI-based ART (Fig 3).

### Impact of switching rates

We calculated levels of NNRTI resistance for 2035 for different average switching delays and percentages of women eligible for DTG-based ART. We considered the effect of a modified switching rate both in suppressed individuals (Fig 4A) and in individuals on a failing regimen (Fig 4B). The predicted levels of NNRTI resistance range from 14.5% to 48.8%. The results indicate potential benefits of both strategies to reduce NNRTI resistance. However, as shown by the greater variation in the prevalence of NNRTI resistance in the vertical than horizontal direction in Fig 4, allowing a higher proportion of women access to DTG-based ART has a greater impact than increasing switching rates.

**Figure 4.**
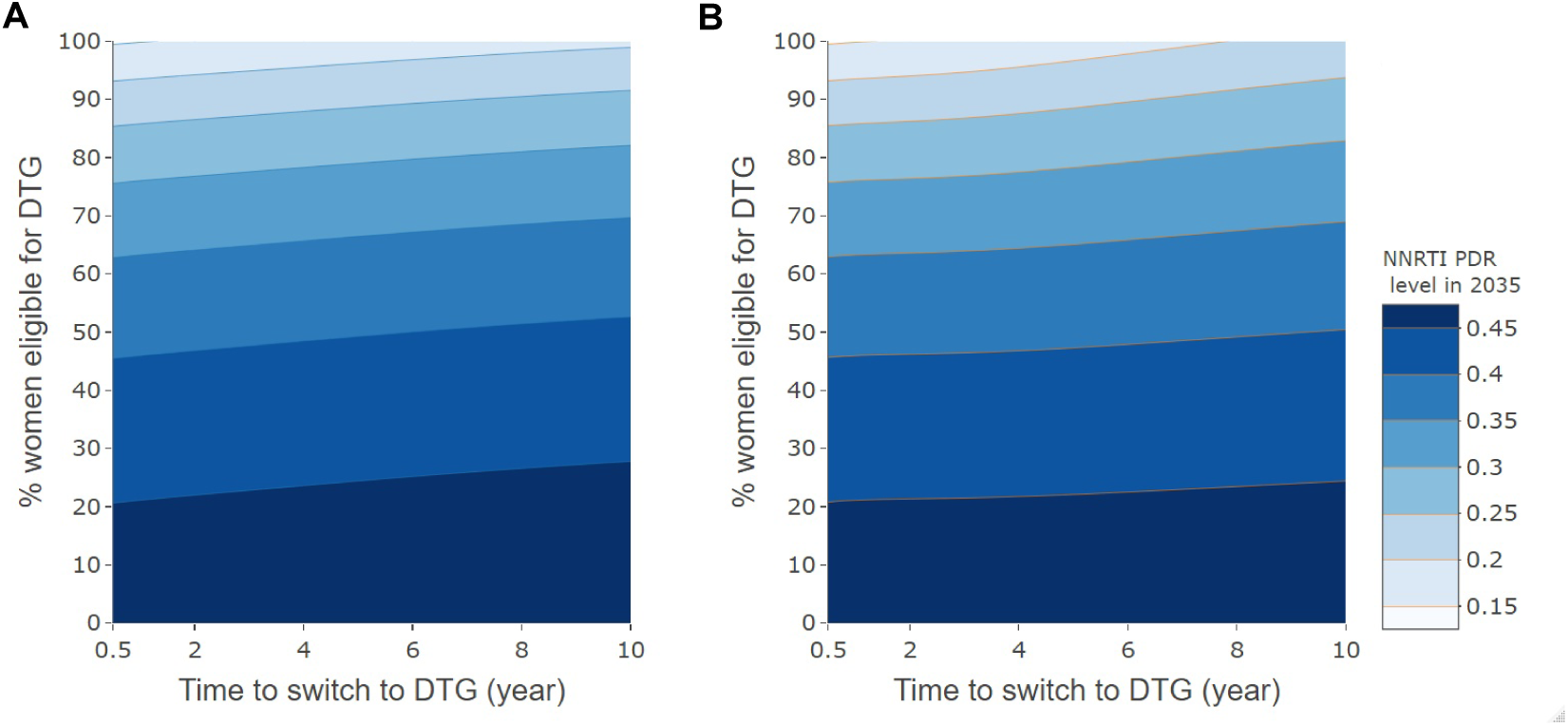
Level of NNRTI pre-treatment drug resistance in 2035, by rate of switching to DTG-based ART and percent women eligible for DTG-based ART. Panel A relates to patients on first-line ART with suppressed HIV-1 replication, and panel B to individuals failing NNRTI-based ART. The average time to switching (i.e. the inverse of the switching rate) varies from 0.5 to 10 years for individuals with viral suppression (panel A) or failure (panel B).

### Impact of DTG-eligibility on the rate of NNRTI failure

As expected from their effect on NNRTI resistance, the different scenarios of the rollout of DTG-based ART also influence the rate of virological failure in women NNRTI-based ART among DTG-ineligible. We find that the proportion of NNRTI failure is increasing over calendar time of ART start for all scenarios, as a result of the increase of NNRTI resistance among NNRTI-initiators (see Fig 5, 2025 vs 2035). In the absence of DTG introduction, we observe a high level of failure in women starting NNRTI in 2035, reaching 42.4% after 2 years of ART. roviding all men with DTG would help diminish the level of failure by 2 years to 32.3% in 2035. This percentage decreases to 28.7%, when including all women not at risk of pregnancy, and to 25.5% if all men and 99% of women are included. Therefore, we find that the increase of virological failure in DTG-ineligible women who still rely on NNRTI can be stopped by the introduction of DTG.

**Figure 5.**
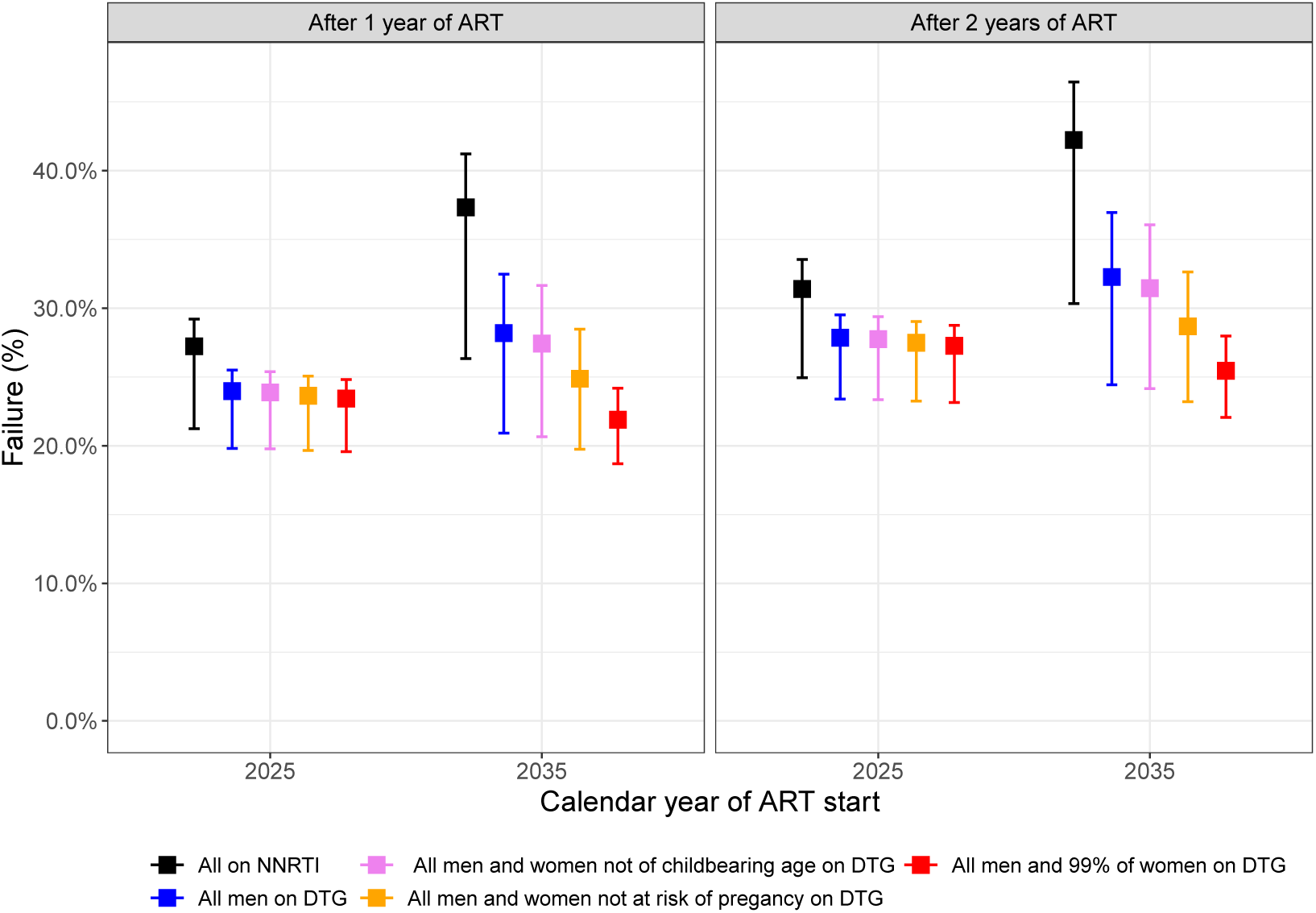
Predicted percentage of women failing NNRTI-based ART after one and two years of ART in 2025 and 2035, depending on the scenario of the rollout of DTG-based ART. Note that the scenario in which DTG is given to all men and 99% of women (in red) replaces the scenario in which DTG was given to all men and women (see “Additional analyses”). Failure is given after 1 and 2 years of ART.

## Discussion

We adapted the epidemiological MARISA model to examine the impact of the scale-up of DTG-based ART on NNRTI pre-treatment drug resistance in South Africa. Overall, our findings suggest that if a large fraction of women is excluded from receiving DTG-based ART, they will not only receive a potentially inferior NNRTI-based regimen but will also face increasing rates of resistance to this regimen due to the population level effects of continued NNRTI use. In contrast, the spread of NNRTI-resistance can be slowed down if DTG-based ART is made accessible both to women at low risk of pregnancy and to people currently on a NNRTI-based first-line regimen, thereby indirectly protecting those still requiring a NNRTI-based treatment. Model simulations emphasize the importance starting on or switching a maximum number of women to DTG-based ART: increasing use of DTG-based regimens was the strategy with the greatest potential to curb the spread of NNRTI resistance. The latter strategy will also lower the risk of virologic failure in women who have to rely on NNRTI-based ART in the future.

While some countries, such as South Africa, first considered to limit access to DTG to men, menopausal women, and women using long-term family planning as a potential policy, the new WHO guidelines state that women should not in principle be excluded from DTG-based ART, even women who are at risk of pregnancy or desire to get pregnant. WHO recommends a woman-centered approach where women should be provided with information about benefits and risks to make an informed choice [24, 16]. It is unclear what proportion of women will effectively receive DTG-based ART, as it depends on individual women s decisions. In this context, model simulations are essential in order to assess the impact of the different options proposed and different levels of DTG uptake. A strength of our model is that it deals with the two most significant sources of uncertainty associated with the introduction of DTG, namely DTG uptake in women and the delay in switching people currently on NNRTI regimens. Despite the uncertainty concerning the uptake of DTG in women, it is likely that a proportion of women will continue to rely on NNRTI-based ART. Therefore, even with the rollout of DTG, NNRTI resistance will continue to be relevant for these women. Compared with other modelling work that assessed risks and benefits of DTG introduction (e.g. [15]), our model focused on its indirect, population-level impact on NNRTI resistance. Rather than assigning a level of NNRTI resistance that is fixed over time, HIV care and disease stages (as in [15]), our model considered the dynamic development of NNRTI resistance under relevant scenarios.

Our model also has several limitations. First, real-world data on the efficacy of DTG, especially in resource-limited settings are scarce. Therefore, we conservatively assumed that DTG has a similar efficacy as NNRTI. The effect of higher DTG efficacy, as suggested by several studies [12, 9], is investigated in supplementary analyses (see S1 Text, Section 5.3). Second, predictions of levels of NNRTI resistance over the next twenty years are naturally uncertain, as reflected by the wide sensitivity ranges in Fig 3. However, despite the uncertainty, it is clear that the different strategies of rolling out DTG-based ART influenced the levels of NNRTI resistance. Finally, the MARISA model includes some simplifying assumptions, e.g. we did not model prevention of mother to child transmission (PMTCT), or treatment interruption. However, relaxing some of these assumptions did not drastically change our conclusion (see S1 Text, Section 5).

Another limitation of this study is the fact that the MARISA model does not take into account resistances to neither NRTI nor DTG. In the context of the introduction of DTG-based ART, modelling of NRTI resistance is particularly relevant as individuals starting on DTG as a functional monotherapy due to resistance to both NRTI backbones - tenofovir and lamivudine - experience higher risk of treatment failure [25]. As they are considerably less frequently transmitted] [26] and revert back quickly [27, 28], NRTI resistances might primarily be an issue for ART-experienced individuals and more specifically, in patients failing NNRTI-based regimens, who often exhibit high levels of NRTI resistance [29]. These patients who are on non-suppressive NNRTI-based regimens are expected to switch to DTG, either after identification of treatment failure, following the new WHO guidelines, or blindly [17]. In the context of modelling the DTG rollout, this consideration has two important implications. First, patients currently failing NNRTI-based regimens are expected to have higher DTG failure rates, mainly due to previously acquired NRTI resistance. Second, due to ongoing viral replication and due to pre-existing NRTI resistance, they are at higher risk of accumulating resistance, which may also lead to the emergence of DTG resistance. So far, data on emergence of DTG resistance is primarily available from patients in whom treatment failure was detected relatively early, which may not be the case in African settings [9]. Therefore, to understand risk inherent in the emergence of DTG resistance, adapting the MARISA model by extending its resistance dimension to both NRTI and DTG resistances will be necessary.

## Conclusion

In conclusion, our study indicates that giving access to DTG-based ART to all women not at risk of pregnancy could limit the increase of NNRTI resistance, but even if all women receive DTG-based ART the level of NNRTI resistance will remain above 10% in South Africa. Our model highlights the importance of a rapid switch of patients currently on NNRTI-based to DTG-based ART in order to limit the increase in NNRTI resistance. Women who remain on NNRTI-based ART will indirectly benefit from a high level of DTG uptake due to a reduced risk of virologic failure.

## Data Availability

Some restrictions will apply.

## Supporting information

**S1 Text Supplementary Material**. Section 1: description of the previous MARISA model. Section 2: description of the adapted MARISA model. Section 3: detailed description of the additional and sensitivity analyses, Section 4: Model equations. Section 5: Additional sensitivity analyses.

## Notes

### Competing Interest Statement

HFG has received unrestricted research grants from Gilead Sciences and Roche; fees for data and safety monitoring board membership from Merck; consulting/advisory board membership fees from Gilead Sciences, Sandoz and Mepha.

### Funding Statement

This study was supported by the National Institutes of Health (www.nih.gov) (National
Institute of Allergy and Infectious Diseases and the Eunice Kennedy Shriver National
Institute of Child Health and Human Development; grant number 2U01AI069924 to
ME) and the Swiss National Science Foundation (Grant No. 174281 to ME). RDK was
supported by the Swiss National Science Foundation (Grant number BSSGI0_155851).

### Author Declarations

All relevant ethical guidelines have been followed and any necessary IRB and/or ethics committee approvals have been obtained.

Any clinical trials involved have been registered with an ICMJE-approved registry such as ClinicalTrials.gov and the trial ID is included in the manuscript.

